# Polygenic Risk Scores for Prediction of Breast Cancer Risk in Women of African Ancestry: a Cross-Ancestry Approach

**DOI:** 10.1101/2021.12.16.21266424

**Authors:** Guimin Gao, Fangyuan Zhao, Thomas U. Ahearn, Kathryn L. Lunetta, Melissa A. Troester, Zhaohui Du, Temidayo O. Ogundiran, Oladosu Ojengbede, William Blot, Katherine L. Nathanson, Susan M. Domchek, Barbara Nemesure, Anselm Hennis, Stefan Ambs, Julian McClellan, Mark Nie, Kimberly Bertrand, Gary Zirpoli, Song Yao, Andrew F. Olshan, Jeannette T. Bensen, Elisa V. Bandera, Sarah Nyante, David V. Conti, Michael F. Press, Sue A. Ingles, Esther M. John, Leslie Bernstein, Jennifer J. Hu, Sandra L. Deming-Halverson, Stephen J. Chanock, Regina G. Ziegler, Jorge L. Rodriguez-Gil, Lara E. Sucheston-Campbell, Dale P. Sandler, Jack A. Taylor, Cari M. Kitahara, Katie M. O’Brien, Manjeet K. Bolla, Joe Dennis, Alison M. Dunning, Douglas F. Easton, Kyriaki Michailidou, Paul D.P. Pharoah, Qin Wang, Jonine Figueroa, Richard Biritwum, Ernest Adjei, Seth Wiafe, GBHS Study Team, Christine B. Ambrosone, Wei Zheng, Olufunmilayo I. Olopade, Montserrat García-Closas, Julie R. Palmer, Christopher A. Haiman, Dezheng Huo

## Abstract

Polygenic risk scores (PRSs) are useful to predict breast cancer risk, but the prediction accuracy of existing PRSs in women of African ancestry (AA) remain relatively low. We aim to develop optimal PRSs for prediction of overall and estrogen receptor (ER) subtype-specific breast cancer risk in women of African ancestry. The AA dataset comprised 9,235 cases and 10,184 controls from four genome-wide association study (GWAS) consortia and a GWAS study in Ghana. We randomly divided samples into training and validation sets. Genetic variants were selected by forward stepwise logistic regression or lasso penalized regression in the training set and the corresponding PRSs were evaluated in the validation set. To improve accuracy, we also developed joint PRSs that combined 1) the best PRSs built in the AA training dataset, 2) a previously-developed 313-variant PRS in women of European ancestry, and 3) PRSs using variants that were discovered in previous GWASs in women of European and African ancestry and were nominally significant the training set. For overall breast cancer, the odd ratio (OR) per standard deviation of the joint PRS in the validation set was 1.39 (95%CI: 1.31-1.46) with area under receiver operating characteristic curve (AUC) of 0.590. Compared to women with average risk (40th-60th PRS percentile), women in the top decile of the PRS had a 2.03-fold increased risk (95%CI: 1.68-2.44). For PRSs of ER-positive and ER-negative breast cancer, the AUCs were 0.609 and 0.597, respectively. The proposed PRS can improve prediction of breast cancer risk in women of African ancestry.

**Author Summary:** Polygenic risk scores have been developed to predict breast cancer risk in non-Hispanic white American women, where polygenic risk score combines the effects of multiple single nucleotide polymorphisms. However, reliable polygenic risk scores do not exist for women of African ancestry, including African Americans, African Barbadians, and indigenous Africans. Due to distinct allele frequencies and linkage disequilibrium structures across populations, polygenic risk scores developed in European ancestry populations have an attenuated predictive value when applied to African ancestry populations. In this study, we constructed polygenic risk scores for African ancestry women by using African ancestry datasets. Since the sample sizes of existing African ancestry datasets are much smaller than those from European-ancestry studies, these polygenic risk scores using only African ancestry datasets may have limited accuracy. To increase the prediction accuracy, we constructed joint polygenic risk scores by combining polygenic risk scores trained in African ancestry datasets with polygenic risk scores that were previously developed using a large European ancestry dataset. Results showed that the joint polygenic risk scores could improve prediction of breast cancer risk in women of African ancestry.

## Introduction

Breast cancer is the most common cancer in women in the United States and worldwide. It is a complex genetic disorder caused by high-penetrance genes, multiple common variants, and non-genetic factors. In the last 10 years, genome-wide association studies (GWAS) had identified more than 180 breast cancer susceptibility loci (1-4). A polygenic risk score (PRS) combines the effects of multiple single nucleotide polymorphisms (SNPs) from GWAS and can achieve a degree of risk stratification that is useful for risk-based programs of breast cancer screening and early detection. PRSs have been developed to predict breast cancer risk in non-Hispanic white, Asian, and Latin American women (5-10). Recently, a large study has developed a 313-variant PRS for breast cancer risks in a European ancestry population (5). This PRS model distinguished breast cancer cases from controls (area under receiver operating characteristic curve, AUC = 0.630 overall), with a better discriminating capacity for ER-positive breast cancer (AUC = 0.641) than for ER-negative breast cancer (AUC = 0.601).

African Americans have higher risk of developing early-onset breast cancer and about 40% higher breast cancer mortality than other racial/ethnic groups in the United States (11), so it is very important to have risk-stratified screening in this population, especially for women age 40 to 49 years. Currently, however, reliable PRS models do not exist for women of African ancestry (AA), including native Africans living in Sub-Saharan Africa and Africa diaspora. Most GWASs of breast cancer were conducted in women of European ancestry, and given the distinct allele frequencies and linkage disequilibrium (LD) structures across populations, PRSs developed in European ancestry populations have an attenuated, though statistically significant, predictive value when applied to African ancestry populations (12, 13). Recently, we showed that the 313-variant PRS can only provide moderate discriminating accuracy in AA, with AUC being 0.571, 0.588, and 0.562 for overall, ER-positive, and ER-negative breast cancer, respectively (14).

Since the sample sizes of existing AA datasets are much smaller than those from European-ancestry studies, using only AA data to develop a PRS may have limited accuracy. To increase the prediction accuracy, we adopted the method of Márquez-Luna et al. (15) to develop joint PRSs by combining an optimal PRS trained in women of African ancestry with the 313-variant PRS that were previously developed in women of European ancestry and PRSs built by using significant variants from previous GWAS.

## Results

We have evaluated the three types of PRS methods described in Materials and Methods: 1) PRSs built by using genome-wide data in women of African ancestry (PRS_AFR_), 2) the 313-variant PRS using effect sizes directly from previous European ancestry studies (PRS_EUR_) and PRSs built by using significant variants from previous studies (PRS_PRE_), and 3) the joint and hybrid PRSs (PRS_Joint_). The evaluation was performed in an African ancestry validation dataset (see below).

### PRSs Built by Using African Ancestry Data Only (PRS_AFR_)

We built PRS models using preset p-value thresholds for filtering SNPs and selecting SNPs by a “hard-thresholding” forward stepwise logistic regression and Lasso regression in the African ancestry training set (see Materials and Methods). Table 1 shows the comparison of the performance of these PRS models developed using AA data only and evaluated in independent validation set. Using the forward stepwise regression approach, the prediction accuracy of PRSs increased as the set p value threshold increased from 10^−5^ to 0.1. The accuracy increased only slightly when the set p value cutoff changed from 0.05 to 0.1, while the numbers of SNPs selected for PRSs for three phenotypes increased about 1.6-fold. Therefore, we used the PRS models with a p value threshold of 0.05 for further analysis. The AUC_adj_ of PRS_AFR29569_, PRS_AFR29004.pos_, PRS_AFR28100.neg_ were 0.535, 0.546, and 0.548 for overall, ER-positive and ER-negative breast cancer, respectively. Here, for example, PRS_AFR29569_ denotes the PRS using 29569 SNPs selected by stepwise forward regression in the African ancestry training dataset.

**Table 1.**
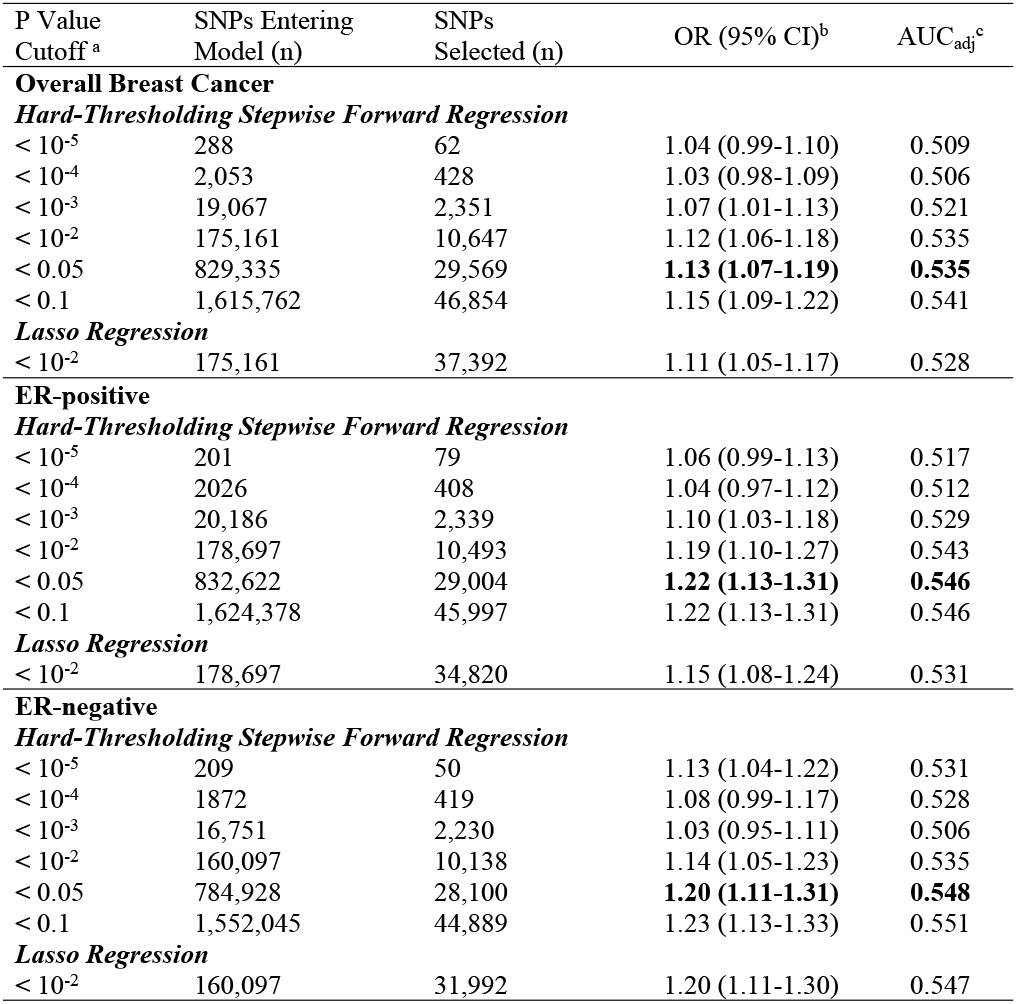
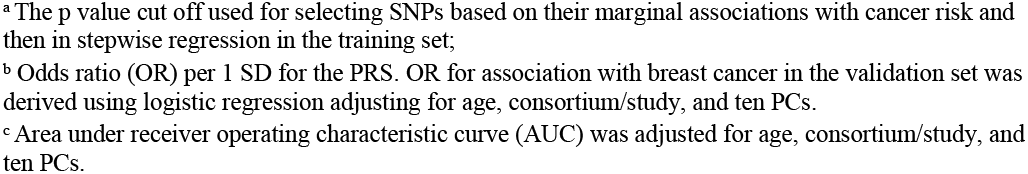
Comparison of the performance of PRS models developed using genome-wide approach in AA data: Results in the validation set

| P Value Cutoff<sup>a</sup> | SNPs Entering Model (n) | SNPs Selected (n) | OR (95% CI)<sup>b</sup> | $AUC_{adj}^c$ |
| --- | --- | --- | --- | --- |
| **Overall Breast Cancer** |  |  |  |  |
| ***Hard-Thresholding Stepwise Forward Regression*** |  |  |  |  |
| $< 10^{-5}$ | 288 | 62 | 1.04 (0.99-1.10) | 0.509 |
| $< 10^{-4}$ | 2,053 | 428 | 1.03 (0.98-1.09) | 0.506 |
| $< 10^{-3}$ | 19,067 | 2,351 | 1.07 (1.01-1.13) | 0.521 |
| $< 10^{-2}$ | 175,161 | 10,647 | 1.12 (1.06-1.18) | 0.535 |
| $< 0.05$ | 829,335 | 29,569 | **1.13 (1.07-1.19)** | **0.535** |
| $< 0.1$ | 1,615,762 | 46,854 | 1.15 (1.09-1.22) | 0.541 |
| ***Lasso Regression*** |  |  |  |  |
| $< 10^{-2}$ | 175,161 | 37,392 | 1.11 (1.05-1.17) | 0.528 |
| **ER-positive** |  |  |  |  |
| ***Hard-Thresholding Stepwise Forward Regression*** |  |  |  |  |
| $< 10^{-5}$ | 201 | 79 | 1.06 (0.99-1.13) | 0.517 |
| $< 10^{-4}$ | 2026 | 408 | 1.04 (0.97-1.12) | 0.512 |
| $< 10^{-3}$ | 20,186 | 2,339 | 1.10 (1.03-1.18) | 0.529 |
| $< 10^{-2}$ | 178,697 | 10,493 | 1.19 (1.10-1.27) | 0.543 |
| $< 0.05$ | 832,622 | 29,004 | **1.22 (1.13-1.31)** | **0.546** |
| $< 0.1$ | 1,624,378 | 45,997 | 1.22 (1.13-1.31) | 0.546 |
| ***Lasso Regression*** |  |  |  |  |
| $< 10^{-2}$ | 178,697 | 34,820 | 1.15 (1.08-1.24) | 0.531 |
| **ER-negative** |  |  |  |  |
| ***Hard-Thresholding Stepwise Forward Regression*** |  |  |  |  |
| $< 10^{-5}$ | 209 | 50 | 1.13 (1.04-1.22) | 0.531 |
| $< 10^{-4}$ | 1872 | 419 | 1.08 (0.99-1.17) | 0.528 |
| $< 10^{-3}$ | 16,751 | 2,230 | 1.03 (0.95-1.11) | 0.506 |
| $< 10^{-2}$ | 160,097 | 10,138 | 1.14 (1.05-1.23) | 0.535 |
| $< 0.05$ | 784,928 | 28,100 | **1.20 (1.11-1.31)** | **0.548** |
| $< 0.1$ | 1,552,045 | 44,889 | 1.23 (1.13-1.33) | 0.551 |
| ***Lasso Regression*** |  |  |  |  |
| $< 10^{-2}$ | 160,097 | 31,992 | 1.20 (1.11-1.30) | 0.547 |

<sup>a</sup> The p value cut off used for selecting SNPs based on their marginal associations with cancer risk and then in stepwise regression in the training set;
<sup>b</sup> Odds ratio (OR) per 1 SD for the PRS. OR for association with breast cancer in the validation set was derived using logistic regression adjusting for age, consortium/study, and ten PCs.
<sup>c</sup> Area under receiver operating characteristic curve (AUC) was adjusted for age, consortium/study, and ten PCs.

Using lasso regression, the best performed PRSs for the three phenotypes were obtained at a p value threshold of 0.01 (Table 1). The prediction accuracy of these best PRSs using lasso were slightly lower than that of the PRSs generated by stepwise regression approach with p value threshold of 0.05, though the lasso PRSs used more SNPs. Therefore, in this study we focused on reporting the PRSs generated by the stepwise approach and the joint PRSs based on these stepwise PRSs.

### The 307-variant PRS (PRS_EUR_) and PRSs Built by Using Variants from Previous Studies (PRS_PRE_)

Directly applying the PRS developed in a European ancestry population (PRS_EUR307_) to our study sample of African ancestry, we found that it was significantly associated with breast cancer risk, with varying prediction accuracy for the three breast cancer phenotypes (Table 2). Of the 322 SNPs identified in previous GWAS, 210, 216, and 174 SNPs had consistent directions of effect for overall, ER-positive, and ER-negative breast cancer in the training set; 58, 47, and 31 were nominally statistically significant for the three phenotypes (Supplemental Table S3). Using effect sizes in the AA training set for these significant SNPs, we calculated a set of recalibrated PRSs (PRS_PRE_). As shown in Table 2, the recalibrated PRS performed worse than the PRS developed in population of European ancestry for overall cancer risk prediction (AUC=0.547 for PRS_PRE58_ vs. AUC=0.574 for PRS_EUR307_) and ER-positive breast cancer (AUC=0.562 for PRS_PRE47.pos_ vs. AUC=0.593 for PRS_EUR307.pos_). However, the recalibrated PRS performed better than the PRS developed in populations of European ancestry for ER-negative breast cancer (AUC=0.575 for PRS_PRE31.neg_ vs. AUC=0.554 for PRS_EUR307.neg_). The AUCs adjusting for age, consortium/study, and ten PCs gave similar results.

**Table 2.** Performance of PRS models using variants selected from previous GWASs: Results in the validation set

|  | No. of SNPs | OR (95% CI)<sup>a</sup> | *P* | AUC | AUC<sub>adj</sub><sup>a</sup> |
| --- | --- | --- | --- | --- | --- |
| **Overall Breast Cancer** |  |  |  |  |  |
| PRS from European ancestry (PRS<sub>EUR307</sub>)<sup>b</sup> | 307 | 1.30 (1.23-1.37) | 2.8x10<sup>-21</sup> | 0.574 | 0.571 |
| PRS from GWASs in European and African ancestry (PRS<sub>PRE58</sub>)<sup>c</sup> | 58 | 1.19 (1.12-1.25) | 5.3x10<sup>-10</sup> | 0.547 | 0.548 |
| **ER-positive** |  |  |  |  |  |
| PRS from European ancestry (PRS<sub>EUR307.pos</sub>)<sup>b</sup> | 307 | 1.43 (1.33-1.53) | 6.1x10<sup>-24</sup> | 0.593 | 0.597 |
| PRS from GWASs in European and African ancestry (PRS<sub>PRE47.pos</sub>)<sup>c</sup> | 47 | 1.26 (1.17-1.35) | 9.2x10<sup>-11</sup> | 0.562 | 0.563 |
| **ER-negative** |  |  |  |  |  |
| PRS from European ancestry (PRS<sub>EUR307.neg</sub>)<sup>b</sup> | 307 | 1.23 (1.13-1.34) | 8.7x10<sup>-7</sup> | 0.554 | 0.557 |
| PRS from GWASs in European and African ancestry (PRS<sub>PRE31.neg</sub>)<sup>c</sup> | 31 | 1.32 (1.21-1.43) | 3.8x10<sup>-11</sup> | 0.575 | 0.578 |

<sup>a</sup> Odds ratio (OR) per 1 SD for the PRS. OR for association with breast cancer in the validation set was derived using logistic regression adjusting for age, study site, and ten PCs. Area under receiver operating characteristic curve (AUC) was adjusted for age, consortium/study, and ten PCs.
<sup>b</sup> For the 313 SNPs reported by Mavaddat et al. (2019)(5) for PRS in women of European ancestry, 307 SNPs appeared in our data of African ancestry.
<sup>c</sup> PRS built from SNPs that were identified in previous GWAS (4, 5, 16, 29) and were significantly replicated ( $p < 0.05$ and the same direction) in the training set of the African ancestry consortia.

As expected, the two types of PRSs built from SNPs identified in previous studies were moderately correlated, with correlation coefficients ranging from 0.427 to 0.521 (Supplementary Table S4). However, these PRSs had almost no correlation with the PRS developed with “hard-thresholding” approach (e.g. PRS_AFR29569_) that used AA data only, suggesting that additional predictive power could be gained if combining these PRSs together.

### The Joint and Hybrid PRS Models

Table 3 shows the prediction performance of the joint and hybrid PRS models (see Materials and Methods) in the validation set. For each phenotype, the three-component joint PRS model performed better than individual PRSs. For overall breast cancer, adding the recalibrated PRS (PRS_PRE58_) to the base model developed using “hard-thresholding” approach (PRS_AFR29569_), the AUC_adj_ increased from 0.535 to 0.558, and further increased to 0.578 after adding the PRS developed in European ancestry population (PRS_EUR307_). Similar results were observed for ER-positive breast cancer. Interestingly, the PRS developed in European ancestry population (PRS_EUR307.pos_) contributed the most to the three-component joint PRS model for ER-positive disease (56%). By contrast, the PRS developed in European ancestry population (PRS_EUR307.neg_) had a small contribution to the three-component joint PRS model for ER-negative breast cancer (19%), while PRS developed using AA data contributed the most. The ORs per unit standard deviation was 1.50 (95% CI: 1.40-1.61) for the joint PRS of ER-positive breast cancer and 1.40 (95% CI: 1.29-1.52) for the joint PRS of ER-negative breast cancer.

**Table 3.** Performance of joint prediction PRS models in the validation set

| | Weight ($\alpha_k$) for<br>each predictor | OR<br>(95% CI)<sup>a</sup> | *P* | AUC<br>(95% CI) | AUC<sub>adj</sub><br>(95% CI)<sup>a</sup> |
| --- | --- | --- | --- | --- | --- |
| **Overall Breast Cancer** |  |  |  |  |  |
| PRS<sub>AFR29569</sub> (genome-wide threshold $P < 0.05$) | | 1.13<br>(1.07-1.19) | $7.8 \times 10^{-06}$ | 0.535<br>(0.520-0.550) | 0.535<br>(0.519-0.551) |
| $\alpha_1$PRS<sub>AFR29569</sub> + $\alpha_2$PRS<sub>PRE58</sub> | $\alpha_1=0.41, \alpha_2=0.59$ | 1.23<br>(1.16-1.30) | $9.3 \times 10^{-14}$ | 0.557<br>(0.542-0.571) | 0.558<br>(0.542-0.575) |
| $\alpha_1$PRS<sub>AFR29569</sub> + $\alpha_2$PRS<sub>PRE58</sub> + $\alpha_3$PRS<sub>EUR307</sub> | $\alpha_1=0.29, \alpha_2=0.11,$<br>$\alpha_3=0.60$ | 1.34<br>(1.27-1.42) | $1.5 \times 10^{-25}$ | 0.579<br>(0.564-0.594) | 0.578<br>(0.563-0.593) |
| PRS<sub>hybrid</sub><sup>b</sup> |  | **1.39**<br>**(1.31-1.46)** | **$2.9 \times 10^{-31}$** | **0.588**<br>**(0.574-0.603)** | **0.590**<br>**(0.575-0.603)** |
| **ER-positive** |  |  |  |  |  |
| PRS<sub>AFR29004.pos</sub> (genome-wide threshold $P < 0.05$) | | 1.22<br>(1.13-1.31) | $2.7 \times 10^{-7}$ | 0.567<br>(0.549-0.586) | 0.546<br>(0.527-0.566) |
| $\alpha_1$PRS<sub>AFR29004.pos</sub> + $\alpha_2$PRS<sub>PRE47.pos</sub> | $\alpha_1=0.45, \alpha_2=0.55$ | 1.33<br>(1.24-1.43) | $3.0 \times 10^{-15}$ | 0.591<br>(0.572-0.609) | 0.578<br>(0.559-0.597) |
| $\alpha_1$PRS<sub>AFR29004.pos</sub> + $\alpha_2$PRS<sub>PRE47.pos</sub> + $\alpha_3$PRS<sub>EUR307.pos</sub> | $\alpha_1=0.31, \alpha_2=0.13,$<br>$\alpha_3=0.56$ | **1.50**<br>**(1.40-1.61)** | **$2.3 \times 10^{-29}$** | **0.616**<br>**(0.598-0.635)** | **0.609**<br>**(0.590-0.629)** |
| **ER-negative** |  |  |  |  |  |
| PRS<sub>AFR28100.neg</sub> (genome-wide threshold $P < 0.05$) | | 1.20<br>(1.11-1.31) | $1.1 \times 10^{-5}$ | 0.546<br>(0.523-0.569) | 0.548<br>(0.525-0.572) |
| $\alpha_1$PRS<sub>AFR28100.neg</sub> + $\alpha_2$PRS<sub>PRE31.neg</sub> | $\alpha_1=0.40, \alpha_2=0.60$ | 1.38<br>(1.28-1.50) | $7.8 \times 10^{-15}$ | 0.592<br>(0.569-0.614) | 0.594<br>(0.571-0.616) |
| $\alpha_1$PRS<sub>AFR28100.neg</sub> + $\alpha_2$PRS<sub>PRE31.neg</sub> + $\alpha_3$PRS<sub>EUR307.neg</sub> | $\alpha_1=0.35, \alpha_2=0.46,$<br>$\alpha_3=0.19$ | **1.40**<br>**(1.29-1.52)** | **$1.1 \times 10^{-15}$** | **0.595**<br>**(0.572-0.617)** | **0.597**<br>**(0.572-0.622)** |

<sup>a</sup> Odds ratio (OR) per 1 SD for the PRS. OR for association with breast cancer was derived using logistic regression adjusting for age, study site, and ten principal components (PCs). Area under receiver operating characteristic curve (AUC) was adjusted for age, study site, and ten PCs.
<sup>b</sup> PRS<sub>hybrid</sub> for overall cancer risk is a linear combination of the joint PRS for ER-positive (PRS<sub>AFR64295</sub> + PRS<sub>PRE47.pos</sub> + PRS<sub>EUR307.pos</sub>) and ER-negative breast cancer (PRS<sub>AFR28100</sub> + PRS<sub>PRE31.neg</sub> + PRS<sub>EUR307.neg</sub>), with weight of 0.62 for ER-positive and 0.38 for ER-negative cancer.

The joint PRS for overall breast had lower prediction accuracy (AUC_adj_ =0.578) than the joint PRSs for ER-positive (AUC_adj_ = 0.609) and ER-negative disease (AUC_adj_ = 0.597). Therefore, we calculated the hybrid PRS for overall breast cancer that combines the PRSs of ER-positive and ER-negative diseases weighted by subtype proportions. The OR per standard deviation of the hybrid PRS was 1.39 (95% CI: 1.31-1.46) with an AUC_adj_ of 0.590. The list of SNPs and corresponding joint effect sizes used for the final joint and hybrid PRSs for the three phenotypes are listed in Supplementary Tables S5, S6, and S7.

The contributing weights of the three components in the joint PRS models (Table 3) were estimated in the validation set, so there might be an overfitting problem. For a robust evaluation of PRS performance, we also used 3-fold cross-validation procedure in the validation set, which is less sensitive to parameter tuning. As shown in Supplementary Figure S1, the AUCs estimated from the cross-validation analysis are similar to the AUCs estimated from analysis using the entire validation set, suggesting the bias due to overfitting is minimal.

Table 4 showed associations between breast cancer risk and percentiles of the joint and hybrid PRSs. Women in the top 10% and 5% of the hybrid PRS had a 2.03-fold (95% CI: 1.68-2.22) and a 2.43-fold (95% CI: 1.92-3.08) elevated overall breast cancer risk compared to women at average risk (PRS in 40th-60th percentiles), respectively. For ER-positive breast cancer, compared to the population average, women in the top 10% and 5% of the joint PRS had a 2.04-fold (95% CI: 1.63-2.57) and a 2.26-fold (95% CI: 1.71-2.99) increased risk, respectively. For ER-negative breast cancer, those in the top 10% and 5% of the joint PRS had a 1.93-fold (95% CI: 1.45-2.56) and a 1.99-fold (95% CI: 1.41-2.82) increased risk, respectively, compared to women at average risk.

**Table 4.** Associations between PRS percentiles and breast cancer risk in the validation set

| PRS Category | No.<br>Control | Overall Breast Cancer |  | ER-positive |  | ER-negative |  |
| --- | --- | --- | --- | --- | --- | --- | --- |
|  |  | No.<br>Case | OR (95% CI) <sup>a</sup> | No.<br>Case | OR (95% CI) <sup>a</sup> | No.<br>Case | OR (95% CI) <sup>a</sup> |
| < 5% | 156 | 77 | 0.65 (0.48-0.88) | 29 | 0.46 (0.30-0.71) | 24 | 0.70 (0.43-1.12) |
| 5% - 10% | 156 | 111 | 0.95 (0.72-1.25) | 38 | 0.66 (0.44-0.97) | 36 | 1.06 (0.70-1.61) |
| 0% - 10% | 312 | 188 | 0.82 (0.66-1.02) | 67 | 0.56 (0.41-0.76) | 60 | 0.88 (0.63-1.23) |
| 10% - 20% | 312 | 182 | 0.78 (0.63-0.98) | 65 | 0.55 (0.40-0.75) | 51 | 0.74 (0.52-1.05) |
| 20% - 40% | 623 | 413 | 0.89 (0.74-1.06) | 178 | 0.72 (0.57-0.90) | 105 | 0.75 (0.57-1.00) |
| 40% - 60% (ref.) | 625 | 460 | 1 (ref.) | 243 | 1 (ref.) | 134 | 1 (ref.) |
| 60% - 80% | 624 | 614 | 1.36 (1.15-1.61) | 268 | 1.09 (0.88-1.35) | 176 | 1.31 (1.02-1.69) |
| 80% - 90% | 312 | 381 | 1.69 (1.39-2.06) | 181 | 1.40 (1.09-1.78) | 124 | 1.90 (1.43-2.53) |
| 90% - 100% | 311 | 464 | 2.03 (1.68-2.44) | 259 | 2.04 (1.63-2.57) | 130 | 1.93 (1.45-2.56) |
| 90% - 95% | 155 | 192 | 1.73 (1.35-2.22) | 117 | 1.83 (1.37-2.45) | 60 | 1.86 (1.30-2.67) |
| >95% | 156 | 272 | 2.43 (1.92-3.08) | 142 | 2.26 (1.71-2.99) | 70 | 1.99 (1.41-2.82) |

<sup>a</sup> Odds ratio (95% confidence intervals) were adjusted for age, consortium and 10 principal components.

The joint and hybrid PRSs were significantly associated with breast cancer risk in women with and without family history of breast cancer (Table 5). We did not see any significant interaction between PRS and family history of breast cancer. In addition, family history was associated with about 1.76 to 2.05-fold increased risk of overall or subtype-specific breast cancer. We only observed slight attenuation of the association of family history with overall breast cancer and ER-negative cancer risk after adjusting for PRS (Table 5).

**Table 5.**
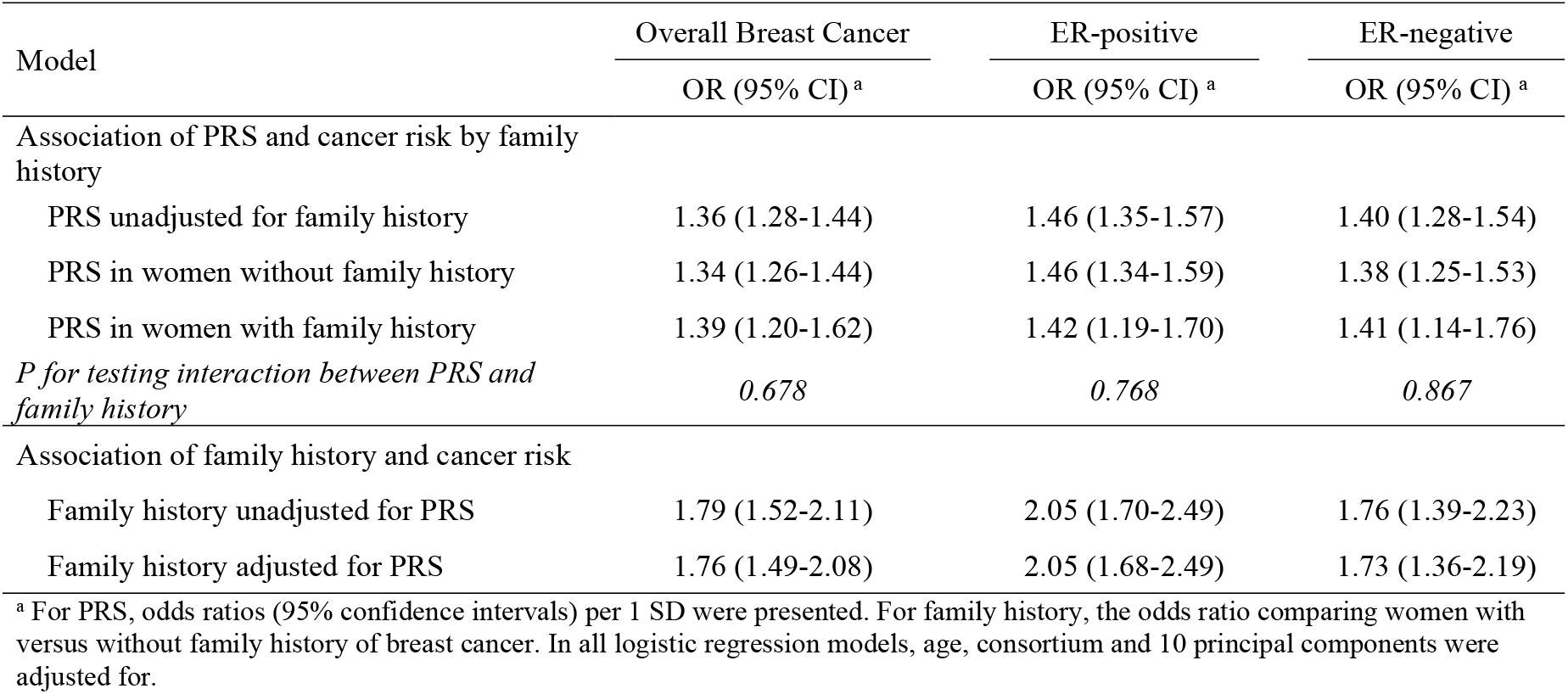
Associations between polygenic risk scores (PRS) and breast cancer risk by family history of breast cancer in the validation set

We did not observe a statistically significant interaction between the joint/hybrid PRSs and age at diagnosis for overall or subtype-specific breast cancer risk (Supplementary Figure S2), although the association between PRS and overall or ER-positive breast cancer risk was weak for women 70 years or older.

We examined association of PRSs and breast cancer risk in two populations: Africans vs. African Americans & African Barbadians. In both populations, PRSs were associated with breast cancer risk and there was no statistically significant interaction (Supplementary Table S8). There was no significant interaction between ancestry groups (<80% African ancestry vs. >80% African ancestry) and PRSs. There was a marginally significant heterogeneity effects of the PRS for overall breast cancer across the five consortium/study, but not for subtype-specific PRSs (Supplementary Figure S3). For overall breast cancer, the PRS has a moderate association in the ROOT and AABC consortia, and a stronger association in the AMBER consortium.

### Absolute Risk of Developing Breast Cancer According to the PRS

Figure 1 shows the estimated life-time and 10-year absolute risks of breast cancer for African Americans according to percentile of the PRSs. The absolute risk of overall breast cancer risk by age 80 years was 19.6% for women in the 99th percentile of the hybrid PRS and 4.1% for women in the lowest 1st percentile. The absolute risk of ER-positive breast cancer by age 80 ranged from 2.2% in the lowest percentile of PRS to 17.8% in the highest percentile of PRS. For ER-negative breast cancer, the absolute risk by age 80 ranged from 1.1% to 5.6%. The dotted line in Figure 1D illustrates the age at which women at different categories of the PRS reach a threshold of 10-year risk of 2%, which corresponds to the average risk for women age 45 years in the U.S. This threshold was reached at 35, 37, and 39 years for women whose PRS is >99th, 95-99th, and 90-95th percentiles, respectively.

**Figure 1.**
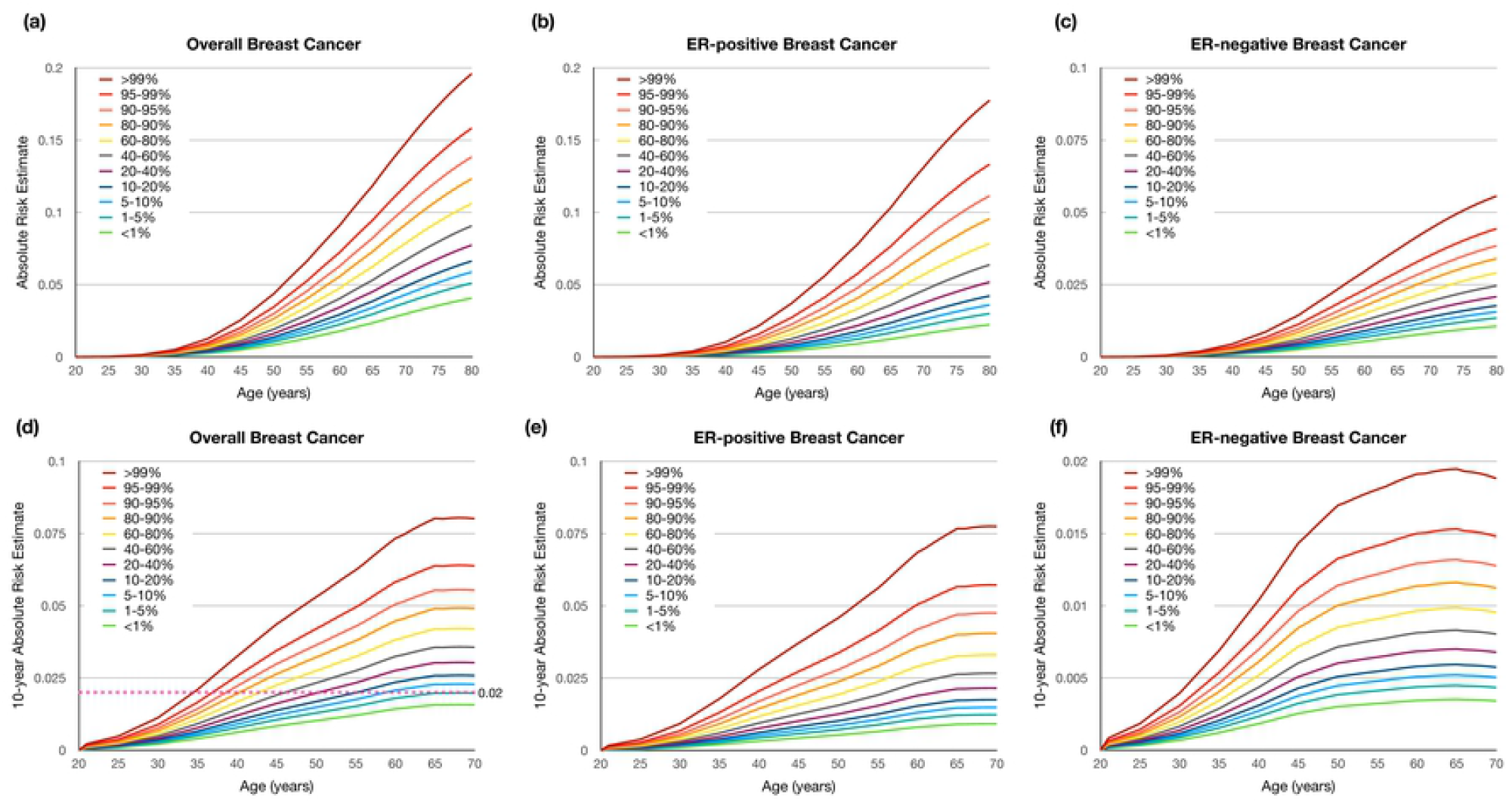
Cumulative life-time and 10-Year Absolute Risk of Developing Breast Cancer

## Discussion

In this study, we developed and validated joint PRSs of breast cancer among women of African ancestry by pooling multiple studies and leveraging an existing polygenic risk score developed in European ancestry population. We adopted the method of Márquez-Luna et al. (15) to develop the joint PRSs that combined 1) the PRS developed with only data from African ancestry, 2) the recalibrated PRS that used variants identified in previous studies and weights estimated in African ancestry training set, and 3) the 313-variant PRS developed in women of European ancestry (5). With AUCs of 0.590, 0.609, and 0.597 for overall, ER-positive, and ER-negative breast cancer, the joint PRSs provide a better predictive value than previous PRS models in African ancestry women. Allman et al evaluated a 77-variant PRS in African Americans and reported an AUC of 0.55 for overall breast cancer risk (12). Wang et al reported an AUC of 0.531 for a 34-variant recalibrated PRS in women of African ancestry (13). Recently, Du et al evaluated the 313-variant PRS using the same dataset as the current study, and reported an AUC of 0.571, 0.588, and 0.562 for overall, ER-positive, and ER-negative breast cancer, respectively (14).

The improved prediction value of the joint PRS models in women of African ancestry may be because it has leveraged the strengths of several PRSs. The 313-variant PRS was developed with very large sample size of 94,075 breast cancer cases and 75,017 controls of European descent (BCAC) (5), so it achieves high precision. The PRS model developed using “hard-thresholding” genome-wide approach in AA datasets has the advantage that the training and validation dataset have the similar LD patterns. The recalibrated PRS utilized variants identified in previous GWAS studies, most of them in European populations, but re-estimated the effect sizes in the African ancestry training set. Of note, the contribution of the individual PRSs to the joint PRSs varied by breast cancer phenotypes. The 313-variant PRS has a better performance in predicting ER-positive than ER-negative breast cancer in both European and African ancestry populations (5, 14). Consistently, it also contributed the most to the ER-positive joint PRS in this study. This may reflect that about 80% of breast cancer is ER-positive disease in breast cancer patients of European ancestry so GWAS data in the BCAC contains more genetic information on ER-positive disease. By contrast, women of African descent patients have higher proportion of ER-negative disease than other populations. Probably because of this, the PRSs trained or recalibrated in our combined AA dataset had the largest contribution to the joint PRS for ER-negative risk.

We also observed that the subtype-specific PRSs performed better than the PRS for overall breast cancer risk. This is probably because of breast cancer etiology heterogeneity and many genetic variants and their effects on ER-positive and ER-negative breast cancers are different (4, 32, 33). Therefore, we generated a hybrid PRS that is weighted average of ER-positive and ER-negative PRSs, and found that prediction accuracy of the hybrid PRS improved moderately. If the finding that “the sum of the parts is greater than the whole” can be confirmed in future studies, it could be a good strategy to estimate omnibus risk of breast cancer (34). While an overall breast cancer risk model and an ER-negative model may be useful for clinical decision making regarding timing and frequency of breast cancer screening, an ER-positive model has the additional advantage of potentially identify high risk women who may benefit from chemoprevention with endocrine agents.

Although the joint PRS models have a better predictive performance than previous PRS models in African ancestry women, the prediction accuracy is still lower than models reported for other racial/ethnic populations. Mavaddat et al reported AUCs of 0.63 and 0.64 for their 313-variant and 3820-variant PRSs, respectively, for predicting overall breast cancer in women of European ancestry (5). Shieh et al examined the performance of 71- and 180-variant PRS for overall breast cancer in a large Latino study and reported AUCs of 0.61 to 0.63 (10). Wen et al examined a 67-variant PRS for overall breast cancer in East Asians and reported an AUC of 0.61 (9). In another PRS study of Asians, Ho et al examined a 287-variant PRS and reported an AUC of 0.613 for overall breast cancer (35). The weaker performance of PRS in women of African ancestry has been observed in other disease phenotypes (36). One study found that the prediction accuracy was 4.9-fold lower in Africans on average compared with that in European populations for 17 phenotypes, while the reduction in accuracy was 1.6-fold in Hispanic/Latino Americans, 1.7-fold in South Asians, 2.5-fold in East Asians (36). These observations are consistent with previous studies which showed that poorer PRS performance is related to genetic divergences between training and target populations (37, 38). Therefore, several factors could account for this disparity, including relatively limited sample size, different LD patterns, allele frequencies, and possible heterogeneity in effect sizes between populations.

To further improve prediction accuracy of PRS in women of African ancestry, it is important to include more racially/ethnically diverse individuals in medical genomic research. The ongoing Confluence project led by U.S. National Cancer Institute has prioritized large-scale genotyping for diverse populations (https://dceg.cancer.gov/research/cancer-types/breast-cancer/confluence-project), so it could improve the prediction accuracy of breast cancer PRS. Advances in methodologies in statistical genetics could also help to develop a better PRS utilizing information hidden in the existing GWAS datasets. For example, sophisticated methods that integrate additional biological information, genetic architecture, and LD information can be promising to apply to diverse populations (39-41). For African Americans, an admixed population, local ancestry could also be tapped to gain statistical power to improve accuracy of genetic risk prediction (42-44).

The AUC, a discriminating accuracy metric, of the new PRS model is moderate, but the model could still provide meaningful risk stratification in the population. Women in the top 5^th^ percentile of the new PRS have more than 2-fold elevated breast cancer risk compared to women at average risk. For women at average risk, the American Cancer Society strongly recommends to initiate regular screening mammography at age 45 years, whose 10-year risk of developing breast cancer is about 2% (45). Based on the PRS, we estimated that about 10% of African American women have 10-year risk of 2% before they reach age 40. These women could start breast cancer screening earlier than age 40 and are possibly eligible for intensive screening programs or chemoprevention trials.

In summary, we proposed joint breast cancer PRSs in women of African ancestry, which has moderate prediction value, but are still not optimal. We found that the joint model can gain more information on ER-positive breast cancer prediction from the existing PRS developed in European ancestry population, while GWAS data from African ancestry contributes more information to the prediction of ER-negative breast cancer.

## Materials and Methods

### Study Participants and Genotyping

This study includes women of African ancestry from four breast cancer GWAS consortia and a study in Ghana, with a combined sample size of 19,419 participants including 9235 breast cancer cases and 10184 controls. Data collection for individual studies of these consortia have been described previously (16-20). Sample size and selected characteristics for each consortium and study are summarized in Supplemental Tables S1. Women in the study sites in United States and Barbados were self-identified as African American or African Barbadian, while women in the African study sites were implied to be of African ancestry. African ancestry was confirmed using GWAS data. For each consortium/study in this project, individual protocols were approved by the relevant Institutional Review Boards at participating centers. All participants provided written informed consent in accordance with the local institutional review boards.

Each consortium/study utilized a different GWAS array. The GWAS of Breast Cancer in the African Diaspora consortium (ROOT) consists of study participants from six studies (16), and samples were genotyped using the Illumina HumanOmni 2.5-8v1 array. After quality control (QC), 1,657 cases (404 ER-positive, 374 ER-negative) and 2,028 controls from the ROOT consortium remained in the analysis. The African American Breast Cancer consortium (AABC) consists of nine epidemiological studies (17, 21, 22). Samples in AABC were genotyped using the Illumina Human 1M-Duo BeadChip. After QC, a total of 3,005 cases (1,517 ER-positive, 986 ER-negative) and 2,713 controls remained in the analysis. The African American Breast Cancer Epidemiology and Risk consortium (AMBER) consists of three studies (18). The AMBER samples were genotyped using the Illumina MEGA array, and after QC, 1406 cases (951 ER-positive, 385 ER-negative) and 2,407 controls remained in analysis. Nine studies with cases and controls of African ancestry contributed samples to the Breast Cancer Association Consortium (BCAC). Genotyping for BCAC was performed using Illumina OncoArray (with 260K GWAS backbone) (23). After removing overlapped samples between BCAC (OncoArray) with AABC, AMBER and ROOT, a total of 2,268 cases (1,127 ER-positive, 613 ER-negative) and 1,406 controls remained for the analysis. The Ghana Breast Health Study (GBHS) includes 899 cases (296 ER-positive, 277 ER-negative) and 1,630 controls (19, 20). Samples in GBHS were genotyped using Illumina Global Screening Array.

### Training Set and Validation Set

In order to pool the samples from these studies, we conducted uniformed imputation using the cosmopolitan reference panel in the 1000 Genomes Project (1KGP) (Phase III release) within each consortium/study by the software IMPUTE2 (http://mathgen.stats.ox.ac.uk/impute/impute_v2.html) (24). After imputation, we filtered in variants (∼15 million SNP or indel) with average minor allele frequency (MAF) > 0.01 and average imputation information score > 0.85. We pooled datasets from the five consortia/study into a combined dataset. Principle component (PC) of genotype data were estimated using EIGENSTRAT in the pooled dataset (25, 26). As shown in the scatter plots of the top five eigenvectors from the principal component analysis (Supplementary Figures S4A and S4B), the first PC can distinguish participants from different continents (Africa vs. North America) and indicates essentially the global proportion of African ancestry. The third and fifth PCs can distinguish countries in Africa. We then randomly split the combined dataset into a training set (n=13,598; 70%) and a validation set (n=5,821; 30%). Model development was conducted in the training set, while the performance of the PRS models were evaluated in the validation set.

### Development of PRSs using Genome-wide Data in Women of African Ancestry

A PRS can be expressed as

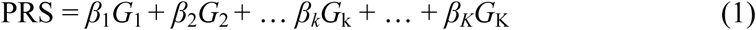

where *β*_*k*_ is the per-allele log odds ratio (OR) for breast cancer associated with SNP *k* and serves as the weight in PRS calculation, *G*_*k*_ is the allele dosage for SNP *k*, and *K* is the total number of SNPs included in the PRS. This form of PRS assumes a log-additive genetic model for individual SNPs, which was considered appropriate in previous PRS development (5-10). To find an optimal PRS, we need to determine which SNPs among all genome-wide variants should be included in the PRS according to association test results from the training dataset, but we do not need to set stringent significance threshold like genome-wide association studies. We used a modified version of the model selection strategy outlined by Mavaddat and colleagues (5), which includes a “hard-thresholding” forward stepwise logistic regression and penalized regression using lasso (27, 28).

First, we performed single SNP-based association tests using multivariable logistic regression in the training set, adjusting for age, consortium/study, and the top ten principal components (PCs). The per allele log-odds ratios or beta coefficients estimated in the single SNP-based analyses are called “marginal” effect sizes. We estimated the association for each of the three phenotypes (overall, ER-positive, and ER-negative breast cancer) in parallel. The model development was also separately for each phenotype, except as otherwise specified (e.g. hybrid model described later).

In the “hard-thresholding” approach, we selected SNPs in three steps. In step 1, we split each chromosome into 5Mb bins and sorted SNPs by p value within each bin. We filtered on linkage disequilibrium such that highly correlated SNPs (LD r^2^ > 0.9) with larger p values were removed. In step 2, we selected SNPs by a series of stepwise forward logistic regression in 5 Mb bin. Only SNPs passing the specified p value thresholds were included in the multivariable models. The SNP with the smallest (conditional) p value was added sequentially to the model, until no further SNPs could be added at the pre-defined threshold. We set p value thresholds to be 10^−5^, 10^−4^, 10^−3^, 10^−2^, 0.05, and 0.1. In step 3, bins of the same chromosome were combined. SNPs on the boundary of two bins (2 Mb boundary) were filtered using LD and stepwise logistic regressions as described in steps 1 and 2. Finally, marginal beta coefficients for all selected SNPs across the genome were compiled together to calculate a PRS according to Equation 1. We labeled this PRS as PRS_AFR_. For low p value threshold (e.g. 10^−4^), we found that it is unnecessary to fit a model including all bins in each chromosome because selected SNPs locate far away from one another and there is no LD. For high p value threshold (e.g. 0.05), there are many (uncorrelated) SNPs on one chromosome and our sample size is limited, so the logistic model including all SNPs cannot be fit reliably.

In the penalized regression using lasso, we first selected SNPs with a given p value threshold and removed SNPs that are in strong LD (r^2^ > 0.9). Then we ran the program glmnet (27) for each chromosome to select SNPs and estimate the corresponding effect sizes. Lastly, we calculated a PRS using the effect sizes of selected SNPs across the genome. We set the p value thresholds to be 10^−5^, 10^−4^, 10^−3^, and 10^−2^ in the lasso regression.

### The 313-variant PRS using Effect Sizes from European Ancestry Population (PRS_EUR_)

The 313-variant PRS was developed by BCAC in a European ancestry population (5). Although its performance in African ancestry populations is not optimal, it still offers moderate discriminatory ability (14). Therefore, we directly applied the weights (beta coefficients) from the 313-variant PRS in the validation set. Of the 313 variants, 6 variants were removed because of low minor allele frequency or imputation score and the remaining 307 variants are shown in Supplemental Tables S1. Here, we use PRS_EUR307_, PRS_EUR307.pos_, and PRS_EUR307.neg_ to denote the PRSs for overall, ER-positive, and ER-negative phenotypes, respectively, where subscript “EUR” indicates the weights are from European ancestry population.

### PRS Using Significant Variants from Previous GWASs and Effect Sizes Estimated in the Training Set (PRS_PRE_)

We postulated that GWAS variants that have consistent directionality in the current study could improve prediction accuracy (13), so we recalibrated the weights for variants discovered from previous GWAS studies in European and African ancestry populations (4, 5, 16). We also examined variants from a recent cross-ancestry GWAS in European and African ancestry populations (29). In particular, we selected variants that were nominally significant (p < 0.05) and had same direction in our AA training set. Of the 322 SNPs, there were 58, 47, and 31 such SNPs for overall, ER-positive, and ER-negative breast cancer, respectively. We used the effect sizes estimated in our AA training set (instead of those from the previous GWASs) as the weights to construct PRS; we labeled these PRSs as PRS_PRE58_, PRS_PRE47.pos_, and PRS_PRE31.neg_, for the three phenotypes. The SNPs used for these PRSs (51, 42, and 25 variants) overlapped with those used in the 313-SNP PRSs described above, but different weights were used in PRS construction.

### Joint and Hybrid PRS Models

To improve risk prediction in diverse populations, Márquez-Luna et al (15) proposed a multiethnic PRS method. The method combines PRS based on European training data with PRS based on training data from the target population (such as African Americans). Márquez-Luna and colleagues showed that the derived multiethnic PRS significantly improve prediction accuracy in the target population and is robust to overfitting (15). Here, we adapted this method to construct a joint PRS as a weighted linear combination of three PRSs:

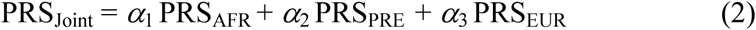

where the weights *α*_1_, *α*_2_, and *α*_3_ were estimated in the validation set using a logistic regression, and PRS_AFR_, PRS_PRE_, and PRS_EUR_ are polygenic risk scores described above. If we let *α*_1_ + *α*_2_ + *α*_3_ = 1, the weights represent the proportional contribution of the three PRSs on the joint PRS.

Since prediction accuracy of the joint PRS for overall breast cancer was relatively low compared to those of the joint PRS for ER-positive and ER-negative breast cancer, we also developed a hybrid PRS as a linear combination of the joint PRSs for ER-positive and for ER-negative breast: PRS_hybrid_ = η PRS_Joint.pos_ + (1 - η) PRS_Joint.neg_, where η = 0.62 was the proportion of ER-positive cases in our study samples.

### Model Evaluation in the Validation Set

For each PRS model described above, we evaluated its performance in the validation set. We calculated the AUC as the measure of the discriminating accuracy of a PRS. We also calculated adjusted AUC (AUC_adj_) using receiver operating characteristic (ROC) regression, in which age, consortium, and the top 10 PCs were adjusted for. To estimate the strength of association, we fit multivariable logistic regression models and calculated odds ratio (OR) and 95% confidence interval (CI) per unit standard deviation of PRS, adjusting for age, consortium, and the top 10 PCs. We also categorized PRSs by percentile (<5%, 5-10%, 10-20%, 20-40%, 40-60%, 60-80%, 80-90%, 90-95%, >95%) in controls, and calculated adjusted OR for each category with 40-60% as the reference group. All analyses were done for overall, ER-positive, and ER-negative breast cancer, separately.

We examined whether age or first-degree family history of breast cancer modified the association between PRS and breast cancer risk by adding interaction terms in logistic regression models. We further examined whether the effect of PRS varied between Africans and African Americans/African Barbadians, between groups defined by African ancestry (<80% vs. >80%), and between the 5 consortium/study.

### Calculation of Absolute Risks

We calculated the lifetime and 10-year absolute risks of developing breast cancer (overall and subtype-specific disease), based on population incidence rates and relative risk estimates for different PRS categories after taking into account the competing risk of dying from causes other than breast cancer, as described previously (6). The theoretical ORs for women in different PRS categories versus women in the 40th-60th percentiles were calculated using the method of Wen et al (9), in which PRS was modeled as continuous predictor of breast cancer risk. Other inputs included age-specific breast cancer incidence rates in African Americans from Surveillance, Epidemiology and End Results (SEER, 2000-2017) (30) and the non-breast cancer mortality rates from Centers for Disease Control and Prevention (CDC 1999-2018) in United States (31). Similarly, we calculated absolute risk of ER-positive and ER-negative breast cancer, using subtype-specific incidence rates from SEER (30) and without accounting for the competing risk of other subtype. Further details are provided in the Supplemental Material and Methods.

We conducted the analyses using R v.3.6.0 and Stata v.16. All tests of statistical significance were two-sided.

## Data Availability

The genotype datasets used in this study are available via dbGaP (https://www.ncbi.nlm.nih.gov/gap/).

https://www.ncbi.nlm.nih.gov/projects/gap/cgi-bin/study.cgi?study_id=phs000851.v1.p1

https://www.ncbi.nlm.nih.gov/projects/gap/cgi-bin/study.cgi?study_id=phs001265.v1.p1

https://www.ncbi.nlm.nih.gov/projects/gap/cgi-bin/study.cgi?study_id=phs000669.v1.p1

https://www.ncbi.nlm.nih.gov/projects/gap/cgi-bin/study.cgi?study_id=phs000383.v1.p1

https://www.ncbi.nlm.nih.gov/projects/gap/cgi-bin/study.cgi?study_id=phs002387.v1.p1

## Acknowledgements

Pathology data of the AMBER project were obtained from numerous state cancer registries (Arizona, California, Colorado, Connecticut, Delaware, District of Columbia, Florida, Georgia, Hawaii, Illinois, Indiana, Kentucky, Louisiana, Maryland, Massachusetts, Michigan, New Jersey, New York, North Carolina, Oklahoma, Pennsylvania, South Carolina, Tennessee, Texas, Virginia). For the studies included in AMBER, individual protocols were approved by the relevant Institutional Review Boards (IRBs) and by the IRBs of participating cancer registries as required. The results reported do not necessarily represent the views of the National Institutes of Health, or the state cancer registries.

GBHS authors acknowledge the research contributions of the Cancer Genomics Research Laboratory for their expertise, execution, and support of this research in the areas of project planning, wet laboratory processing of specimens, and bioinformatics analysis of generated data. The success of this investigation would not have been possible without exceptional teamwork and the diligence of the field staff who oversaw the recruitment, interviews and collection of data from study subjects. Special thanks are due to the following individuals: Korle Bu Teaching Hospital,Accra—Dr Adu-Aryee, Obed Ekpedzor, Angela Kenu, Victoria Okyne, Naomi Oyoe Ohene Oti, Evelyn Tay; Komfo Anoyke Teaching Hospital, Kumasi— Marion Alcpaloo, Bernard Arhin, Emmanuel Asiamah, Isaac Boakye, Samuel Ka-chungu and; Peace and Love Hospital, Kumasi—Samuel Amanama, Emma Abaidoo, Prince Agyapong, Thomas Agyei, Debora Boateng-Ansong, Margaret Frempong, Bridget Nortey Mensah, Richard Opoku, and Kofi Owusu Gyimah. The study was further enhanced by surgical expertise provided by Dr Lisa Newman of the University of Michigan and by pathological expertise provided by Drs. Stephen Hewitt and Petra Lenz of the National Cancer Institute and Dr. Maire A. Duggan from the Cumming School of Medicine, University of Calgary, Canada. Study management assistance was received from Ricardo Diaz, Shelley Niwa, and Usha Singh. Appreciation is also expressed to the many women who agreed to participate in the study and to provide information and biospecimens in hopes of preventing and improving outcomes of breast cancer in Ghana.

## Supporting Information

### Supplementary Authorship

The GBHS Study Team

Florence Dedey^1^, Richard Biritwum^2^, Lawrence Edusei^1^, Verna Vanderpuye^1^, Ernest Adjei^3^, Francis Aitpillah^3^, Joseph Oppong^3^, Margaret Frempong^4^, Jonine Figueroa^5^, Louise Brinton^6^, Thomas U. Ahearn^6^, Ernest Osei-Bonsu^3^, Nicholas Titiloye^3^, Michelle Brotzman^6^, Ann Truelove^7^, Evelyn Tay^1^, Naomi Oyoe Ohene Oti^1^, Victoria Okyne^1^, Isaac Boakye^3^, Bernard Arhin^3^, Marion Alcpaloo^3^, Emma Abaidoo^4^, Prince Agyapong^4^, Joe Nat Clegg-Lamptey^1^, Joel Yarney^1^, Kofi Nyarko^2^, Daniel Ansong^3^, Baffour Awuah^3^, Seth Wiafe^4^, Beatrice Addai Wiafe^4^, Montserrat Garcia-Closas^6^

Affiliations:

1. Korle Bu Teaching Hospital, Accra, Ghana
2. University of Ghana, Accra, Ghana
3. Komfo Anoyke Teaching Hospital, Kumasi, Ghana
4. Peace and Love Hospital, Kumasi, Ghana
5. University of Edinburgh, Edinburgh, Scotland
6. U.S. National Cancer Institute, Bethesda, MD
7. Westat, Inc., MD, USA

### Supplementary Material and Methods (including supplementary Figure S1-4)

**Supplementary Table S1**. Descriptive characteristics of study samples.

**Supplementary Table S2**. Beta coefficients of the 313-variant polygenic risk score model

**Supplementary Table S3**. Beta coefficients estimated in the training set for variants selected from previous GWASs.

**Supplementary Table S4**. Correlation coefficients among selected polygenic risk scores in the validation set.

**Supplementary Table S5**. Beta coefficients for hybrid polygenic risk score for overall breast cancer risk prediction.

**Supplementary Table S6**. Beta coefficients of joint polygenic risk score for ER-positive breast cancer risk prediction.

**Supplementary Table S7**. Beta coefficients of joint polygenic risk score for ER-negative breast cancer risk prediction.

**Supplementary Table S8**. Associations between polygenic risk scores and breast cancer risk by race in the validation set.

**Supplementary Figure S1**. Area under the receiver-operating characteristic curves (AUC) for the 3 polygenic risk score models using the entire validation set or from 1000 3-fold cross-validations.

**Supplementary Figure S2**. Association of the polygenic risk score and breast cancer risk in different age categories (in years) in the validation set.

**Supplementary Figure S3**. Association of the polygenic risk score and breast cancer risk in different consortium/study in the validation set.

**Supplementary Figure S4**. Scatter plots of the top 5 eigenvectors from principal component (PC) analysis according to consortium/study (A) and country (B).

